# Characterization of a novel, low-cost, scalable vaporized hydrogen peroxide system for sterilization of N95 respirators and other COVID-19 related personal protective equipment

**DOI:** 10.1101/2020.06.24.20139436

**Authors:** Nikhil Dave, Katie Sue Pascavis, John Patterson, David Wallace, Abhik Chowdhury, Morteza Abbaszadegan, Absar Alum, Pierre Herckes, Zhaobo Zhang, Michael Kozicki, Erica Forzani, S. Jimena Mora, Josh Chang, Clinton Ewell, Tyler Smith, Mark Naufel

## Abstract

Due to the virulence of severe acute respiratory syndrome coronavirus 2 (SARS-CoV-2), the pathogen responsible for the respiratory disease termed COVID-19, there has been a significant increase in demand for surgical masks and N95 respirators in medical clinics as well as within communities operating during the COVID-19 epidemic. Thus, community members, business owners, and even medical personnel have resorted to alternative methods for sterilizing face coverings and N95 respirators for reuse. While significant work has shown that vaporized hydrogen peroxide (VHP) can be used to sterilize N95 respirators, the cost and installation time for these sterilization systems limit their accessibility. To this end, we have designed and constructed a novel, cost-effective, and scalable VHP system that can be used to sterilize N95 respirators and other face coverings for clinical and community applications. N95 respirators inoculated with P22 bacteriophage showed a greater than 6-log_10_ reduction in viral load when sterilized in the VHP system for one 60-minute cycle. Further, N95 respirators treated with 20 cycles in this VHP system showed comparable filtration efficiency to untreated N95 respirators in a 50 to 200 nanometer particulate challenge filtration test. While a 23% average increase in water droplet roll-off time was observed for N95 respirators treated with 5 cycles in the sterilization, no breakdown in fluid resistance was detected. These data suggest that our VHP system is effective in sterilizing N95 respirators and other polypropylene masks for reuse. Relating to the present epidemic, deployment of this system reduces the risk of COVID-19 community transmission while conserving monetary resources otherwise spent on the continuous purchase of disposable N95 respirators and other face coverings. In summary, this novel, scientifically validated sterilization system can be easily built at a low cost and implemented in a wide range of settings.

## Introduction

The COVID-19 outbreak has resulted in an unprecedented strain on the global personal protective equipment (PPE) supply chain [1]. Particularly, the demand for face coverings such as surgical masks and medical-grade N95 respirators has increased substantially throughout the COVID-19 pandemic in order to prevent the spread of SARS-CoV-2 [2, 3]. In turn, medical professionals and community members have turned to alternative mechanisms for sterilizing and reusing their existing stockpile of face coverings and N95 respirators. Given the prevalence of SARS-CoV-2 and its impact on public health, the terms “sterilization of viral loads” and “sterilization” will be used interchangeably in this work.

Recent literature highlights the efficacy of large-scale vaporized hydrogen peroxide (VHP) systems to sterilize N95 masks for reuse. The Food and Drug Administration (FDA) has reported achieving sterilization of N95 respirators using a Battelle VHP system, as indicated by 6-log_10_ reduction of bacteria, without affecting the filtration efficiency [4,5]. The N95 fit was unaffected after 20 treatment cycles and the elastic did not degrade within 30 treatment cycles [4,5]. Additionally, Duke University validated that proper sterilization was achieved, while Yale University and University of Manitoba, Winnipeg Canada demonstrated eradication of SARS-Cov-2 and other viruses acting as proxies [6,7,8]. Further, sterilization of disposable N95 masks using VHP has been approved in some cases by both the FDA and the CDC, citing its biocidal activity against various biological particles including bacteria, severe acute respiratory syndrome coronavirus 2 (SARS-CoV-2), and other viruses [9].

However, these systems, including the Bioquell BQ-50 device and similar machines, are high in cost and require significant specialized labor to install, making it difficult to scale this model to the needs of individual community members [10]. Given the increasing importance of PPE both in health care facilities as well as in various communities as they reopen in the coming months, we sought to develop a cost-effective and scalable device that can sterilize any type of face covering, including N95 respirators.

Unlike other commercially available systems, our vaporized hydrogen peroxide system operates at reduced pressure, which causes the feed solution of hydrogen peroxide in water to vaporize and occupy the vessel in the gas phase, based on the principles of Raoult’s law and Dalton’s law of partial pressures [11]. The pressure within a vacuum chamber is reduced, causing a solution of hydrogen peroxide and water to vaporize and occupy the vessel in the gas phase. By controlling the absolute pressure within the chamber, the relative concentration of vaporized hydrogen peroxide compared to other gases, such as water vapor, oxygen, and nitrogen, can be controlled. Raoult’s law also dictates that the partial pressure of the hydrogen peroxide vapor can be controlled with the concentration of the hydrogen peroxide feedstock. Our system operates using a 3% hydrogen peroxide solution at room temperature.

In this system configuration, an industrially-available vacuum chamber is used in conjunction with an HVAC-type rotary-vane vacuum pump to evacuate the chamber for VHP treatment. A nonreactive fiber matrix, in this case woven fiberglass cloth, accelerates the vaporization of hydrogen peroxide solution by providing surface area and wicking, which facilitates exposure of the hydrogen peroxide solution to the chamber atmosphere. The use of material that will not react with hydrogen peroxide prevents the consumption or explosive decomposition of hydrogen peroxide [12]. One or more dividing plates are used in order to separate the treated items, ensuring adequate gas diffusion (see Supporting Information for a comprehensive overview of the construction and operation of the Luminosity Lab’s VHP system).

Given the low cost and scalability of our VHP sterilization system, we sought to validate this system for clinical and community use by demonstrating efficacy for sterilization of surgical N95 respirators due to their widespread use and well-defined standards for minimum performance as dictated by the FDA and NIOSH. Validating a system for sterilization of N95s should involve not only biological validation but also consideration of the effects of sterilization on standards such as fit, inhalation resistance, and most importantly an N95’s required filtration efficiency of 95% or higher for non-oily solid and liquid aerosols [13, 14, 15]. Furthermore, any surgical mask such as a surgical N95 must be validated for fluid-resistant performance as required by the FDA [14]. A sterilization system used by healthcare facilities and certain industries and communities should not allow the performance of N95s to fall below FDA and NIOSH guidelines [13]. Based on these criteria, we experimentally validated the system’s virucidal capability, impact on the filtration efficiency of N95 respirators, and effect on N95 respirator hydrophobicity.

## Methods

### Analytical derivation of vaporized hydrogen peroxide concentration

The concentration of VHP directly affects the biocidal capabilities of a VHP sterilization system. Thus, we determined the VHP concentration inside the system both analytically and experimentally. Raoult’s law was used to determine the partial pressure of VHP in the vacuum chamber as a function of total chamber pressure and concentration of hydrogen peroxide feedstock solution. Raoult’s law can be expressed as in (1):

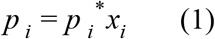

where *p* _*I*_ is the vapor pressure of a single compound in a mixture of liquids at a given temperature, *p* _*i*_* is the vapor pressure of that compound in its pure form, and *x*_*i*_ is the mole fraction of the compound in the solution. If both the vapor pressure at a given temperature of pure hydrogen peroxide and the mole fraction of hydrogen peroxide in the hydrogen peroxide solution are known, the partial pressure of hydrogen peroxide in the gas phase can be calculated. As a result, the concentration in parts per million (ppm) of hydrogen peroxide in the gas phase can be determined if the total pressure of gases in the chamber is known.

The mole fraction of hydrogen peroxide in aqueous solution is found as in (2).

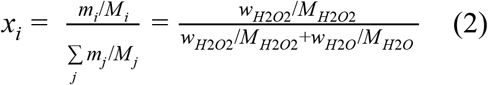

where *w*_*H*2*O*2_ is the mass fraction of hydrogen peroxide in the solution, *w*_*H*2*O*_ is the mass fraction of water in the solution, *M*_*H*2*O*2_ is the molar mass of hydrogen peroxide, and *M*_*H*2*O*_ is the molar mass of water.

From the periodic table of the elements, it can be determined that the molar mass of water is 18.02 g/mol and the molar mass of hydrogen peroxide is 34.01 g/mol.

For a 3% weight fraction solution of hydrogen peroxide, the mole fraction is found in (3).

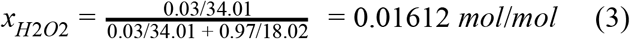

In the case of the system described, the vapor pressure of pure hydrogen peroxide at 25°C is given as 2.1 torr by Maass [16]. It can therefore be established that based on (1), the partial pressure of hydrogen peroxide in a 3% weight fraction solution at 25°C is 0.03385 torr as in (4):

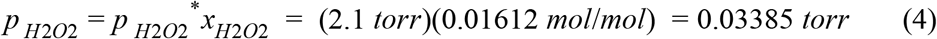

The gas concentration can be expressed as the ratio of the number of particles of the gas of interest to the number of particles of all gases in the container, or equivalently the ratio of the partial pressure of the gas of interest to the total pressure of all gases in the container. This concentration may be expressed in parts per million (ppm) as in (5):

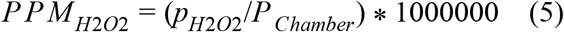

If the target concentration of VHP is given as 1200 ppm 4], the absolute pressure of all gases in the vacuum chamber is therefore 28.21 mmHg, as in (7):

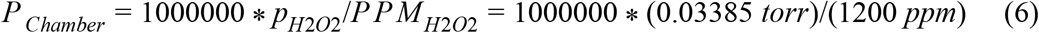

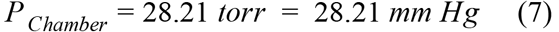

### Experimental validation of vaporized hydrogen peroxide concentration

Hydrogen peroxide (H_2_O_2_) 50% weight fraction, potassium permanganate (KMnO_4_), sodium oxalate, and sulfuric acid were purchased from Sigma Aldrich. The solutions were prepared using MilliQ water. High-level peroxide test strips were obtained from Bartovation. All the solutions were kept in light-blocking caramel containers tightly closed in the dark at a temperature between 2 to 8 °C. Relative pressure was measured with a digital manometer Dwyer series 475 mark III. The colorimetric absorption change was measured by an Arizona State University-developed iOS application installed on an Apple iPhone 6, which provides high quality imaging hardware and software.

All solutions, including commercially available H_2_O_2_ 3% weight fraction, were titrated by the well-known standard redox method with KMnO_4_, previously standardized with sodium oxalate. Each solution was acidified with concentrated sulfuric acid.

The same procedure previously described was followed. For all the tests, 40 mL of solution was used in a chamber of 18 L at room temperature. 40 mL was selected to ensure adequate solution was present in the chamber to prevent total evaporation of solution, but other amounts may be used if it is ensured that the solution does not evaporate completely during system operation. The chamber was closed after 5 minutes of maximum vacuum and treated for 1 hour. The testing configuration described herein is pictured in Fig. 1.

**Fig. 1.**
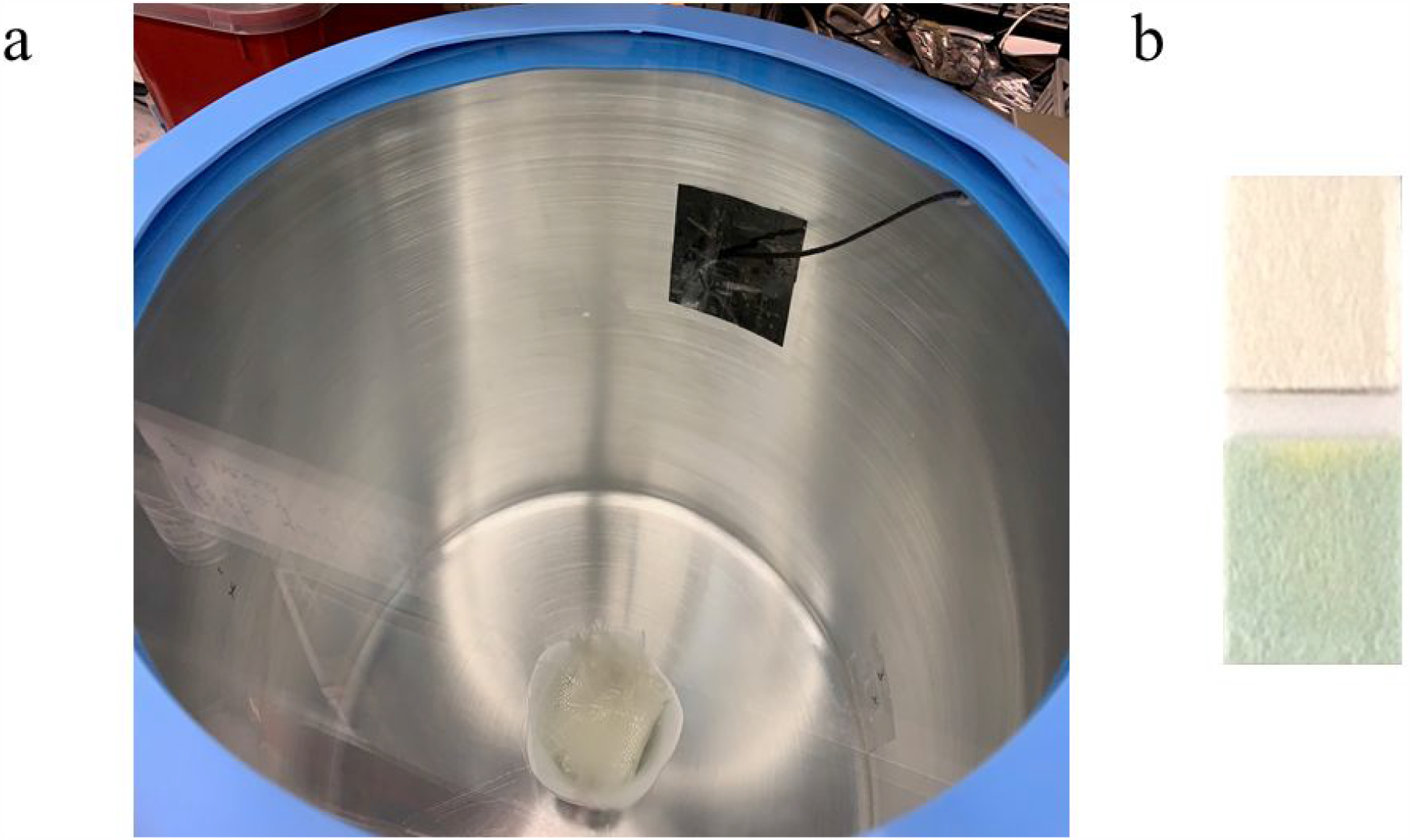
a. System designed for measurement of H_2_O_2_ concentration inside the chamber. b. Color of the hydrogen peroxide test strip corresponding to the unexposed reference sample (top) and the exposed post-treatment sample with H_2_O_2_ 3% weight fraction (bottom).

The strips were dipped into MilliQ water for 2 seconds, removed, and shaken to eliminate excess water. Three strips were used per test. A patch was designed with aluminized Mylar film, a gas barrier, to cover the strips inside the chamber attached with double sided tape. After one hour a cord was pulled allowing the exposure of the strips to the vapor for 30 seconds. Before re-introducing air in the chamber, the Relative Pressure (P_rel_) was measured. The pictures were taken immediately after opening the chamber. The app allows three pictures per strip, having as reference a strip with no reaction or previous treatment.

The data is obtained from our laboratory-custom apparatus in a comma-separated values (csv) file with the corresponding red, green and blue (RGB) intensities for the reference and sensor strip, and absorbance for each component is also provided [17, 18]. The calibration curve was plotted as the difference between the average of the red absorbance for the three strips at the end of the experiment and the corresponding to 0 ppm (MilliQ water). The H_2_O_2_ vapor concentrations (x_H2O2, v_ and ppm_v_ concentration) were calculated by Dalton’s Law considering the chamber’s absolute total pressure (calculated from a measured differential pressure by a gauge the atmospheric pressure), and the H_2_O_2_ partial pressure, previously calculated from literature data [16] containing information of H_2_O_2_ equilibrium partition and the liquid H_2_O_2_ molar fraction (x_H2O2, l_) calculated from the molarity determinate by titration.

After each trial, the chamber was washed with water and allowed to dry and ventilate in a fume hood for 20 min.

### Validation of virucidal activity using P22 bacteriophages as a proxy for SARS-CoV-2 Viral Preparation and Assay

Bacteriophage P22 (ATCC® 19585-B1™) was propagated using *Salmonella typhimurium* (ATCC® 19585™) as the host bacterium using the double agar layer (DAL) technique. Briefly, 1 mL of the sample and 1 mL of the host cell bacteria, in the log-phase of growth, were added to a 5 mL of melted top agar in a test tube which was kept in a water bath at 48°C. The mixture was gently poured onto a bottom agar plate and kept undisturbed to let the top agar to solidify. Then, the plate was incubated upside down at 37°C, and plaques were counted after 24 hours of incubation. A positive and a negative control was included in every DAL assay.

The bottom agar plates were prepared using Tryptic Soy Agar (TSA) (Difco Laboratories, Division of Becton Dickinson & Co.). Prepared TSA was sterilized using an autoclave, cooled to 55°C and then dispensed into petri plates (20 ml per plate). TSA in plates were allowed to solidify and then stored at 4°C until used.

The top agar was prepared using Tryptic Soy Broth (TSB) (Difco) by adding 0.7% agar (Difco). Five mL of the top agar medium were dispensed to test tubes, capped and then autoclaved. The top agar tubes were stored at 4°C until used.

### Inoculation and Test Procedure

A surgical 3M 1860 N95 mask was cut to 2.5 × 2.5 cm pieces using a sterilized pair of scissors to obtain test coupons. Three coupons were placed in a sterilized petri dish, and each was inoculated with P22 bacteriophages at a total concentration of 1.6×10^8^ PFUs per coupon by transferring 100 µL of the stock solution to their inner surface. The inner surface of the mask was selected based on its higher absorbance capability compared to the outer surface which is highly hydrophobic (data not shown). The inoculated coupons were allowed to dry at room temperature for 5 minutes to equilibrate with the mask material.

Triplicate sets of inoculated coupons were placed in the Ozone generating, and the Vaporized Hydrogen Peroxide sterilization systems. Both systems were operated according to the parameters specified earlier.

After the operation cycle completed, test coupons were retrieved from the systems. Each coupon was placed in a 50 mL tube containing 30 mL of elution buffer. The tube was vortexed to recover viruses from the coupon, and then the recovered buffer was analyzed using DAL technique.

### Validation of N95 respirator filtration efficiency before and after treatment

The capture efficiency tests were performed using a custom set-up. Challenge aerosols were generated using a medical nebulizer (drive medical, Port Washington, NY, USA) from aqueous solutions. Tests were performed using fumed silica nanoparticle slurry solutions which have been extensively characterized [19]. The nebulization resulted in a broad challenge aerosol distribution and typical observational range was from 50-200 nm, a range which incorporates particles smaller than individual virions. This challenge aerosol covers the range of aerosols used in the NIOSH tests methods, 75 ± 20 nm NaCl particles for N type masks [20] and dioctyl phthalate (185 ± 20 nm) particles for P99 masks [21].

The challenge aerosol was passed through a trap bottle to remove larger particles. The aerosol then was measured directly or passed through a 25 mm diameter punch sample of the mask material, held in a filter cassette. The size resolved particle number concentration were determined with a Scanning Mobility Particle Sizer (SMPS) set-up consisting of a TSI 3088 Soft X-Ray neutralizer, a TSI 3082 aerosol classifier, a TSI 3085A Nano differential mobility analyzer (DMA) and a TSI 3752 high concentration condensation particle counter (CPC) (TSI, Shoreview, MN, USA). All tests were run in triplicate and for at least 10 minutes each. The respirators used for validation were surgical 3M 1860 N95s.

### Validation of N95 respirator fluid resistance before and after treatment

The fluid resistance of the N95 mask surface of control-group (untreated) and experimental-group (5 cycles of VHP treatment) N95 masks was quantified by means of a water drop deflection test. Distilled water was dropped from a fixed height of 17 cm above the base of the mask onto the surface of a mask. The mask was tilted such that the nose-clip edge of the mask was elevated 5 cm above the base of the mask in order to ensure successful roll-off of drops from the mask surface. A video camera was then used to record the falling drops of water, and the amount of time taken by the drops to traverse the surface of the mask and reach the bottom of the mask was quantified and compared between the control-group masks and the experimental-group masks. Each mask was tested with three different water drops, and three of each type of mask (control and experimental) were used to provide a total of 18 test results for comparison. In addition to the quantitative observations provided by the video recording, it was also qualitatively noted whether any absorption of the water drops into the mask surface was observable. The respirators used for validation were surgical 3M 1860 N95s.

## Results

### Experimental validation of vaporized hydrogen peroxide concentration in the system

A method using commercial peroxide sensor strips with a reaction based on peroxidase and a chromophore compound developing color in the blue-cyan spectrum region was optimized for assessing the concentration of VHP as described in the experimental section. The sensor strips for liquid solutions were adapted and used with our iOS application. Given the reaction chromophore color and comparison with light component absorbance for green, blue and red, we determined the red light component was the most sensitive to use for correlation with the H_2_O_2_ vapor concentration. Fig. 2 shows the calibration curve obtained in the chamber with sensitivity of 8 × 10^−5^ a.u. / ppm and an R-squared correlation coefficient (R^2^) of 0.985. Given this high sensor performance correlation, we were able to determine the H_2_O_2_ vapor concentration under normal system operation conditions for the chamber using a 3% weight fraction H_2_O_2_ solution, which was previously titrated and measured by duplicate as shown in Table 1. This solution provided an operational vapor H_2_O_2_ concentration of 573 ± 107 (SD) ppm.

**Fig. 2.**
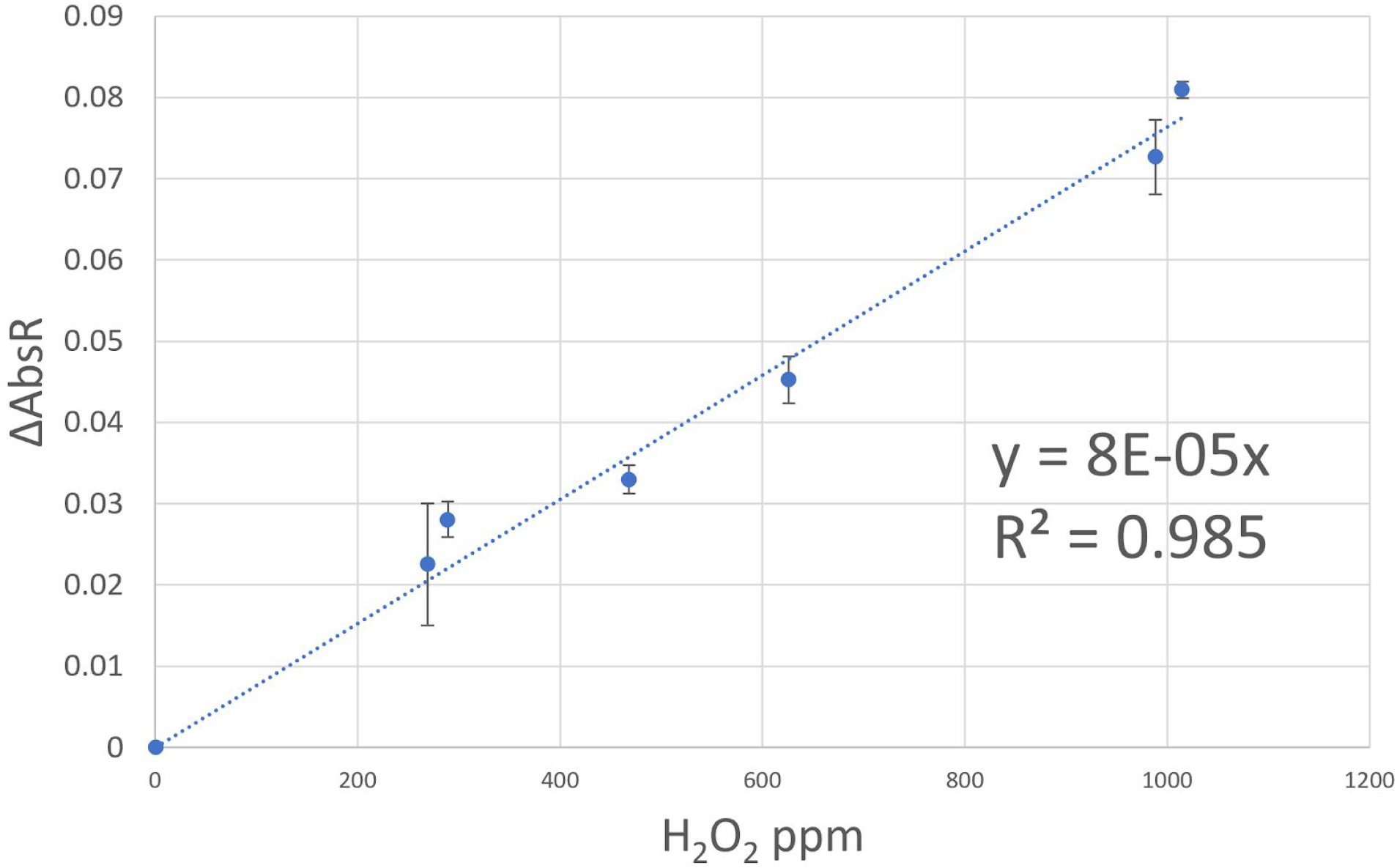
Calibration curve for H_2_O_2_ solutions 0-5%. Error bars represent the standard deviation of the red light component absorbance.

**Table 1.** Results from experiments run for commercially available H_2_O_2_ 3% weight fraction solution.

| | $P_{rel}^a$ | $P_{atm}^a$ | $P_{Abs}^b$ | $X_{H_2O_2}$ | Pp | ppm | AbsR<sup>c</sup> | ΔAbsR | $C_{H_2O_2, v}$ (ppm) |
| --- | --- | --- | --- | --- | --- | --- | --- | --- | --- |
| **Test 1** | -0.935 | 1.014 | 60 | 0.0167 | 0.035 | 582 | 0.047802 | 0.037256 | 466 |
| **Test 2** | -0.929 | 1.0098 | 60.6 | 0.0167 | 0.0349 | 576 | 0.054483 | 0.043937 | 681 |

$P$ in bar units; <sup>b</sup> $P$ in torr units; <sup>c</sup> average of the AbsR (a.u.)

### Validation of virucidal activity using P22 bacteriophages as a surrogate for SARS-CoV-2

The viral inactivation data is presented in Table 2. The vaporized hydrogen peroxide system achieved sterilization of viruses on N95 mask coupons. In all tests, greater than 6-log_10_ reductions of P22 bacteriophage were achieved under the test conditions, which surpasses requirements for sterilization [5, 22]. In this experiment, the test samples were exposed to the equilibrium VHP atmosphere within the chamber for a total time of 60 minutes. The observed 6-log_10_ reduction of the viral proxy–a similar result to Yale University’s validation of the BQ-50 VHP system (Bioquell, Horsham, PA) and NIOSH and CDC’s evaluation of UVGI for sterilization of N95s–demonstrates the system’s effectiveness against SARS-CoV-2 [7, 13].

**Table 2.** Inactivation of P22 bacteriophages on N95 mask coupons using VHP sterilization system.

| **Treatment** | **Replicate** | **Initial**<br>**Log<sub>10</sub> Inoculated** | **Final**<br>**Log<sub>10</sub> Recovered** | **Log<sub>10</sub>**<br>**Reduction** |
| --- | --- | --- | --- | --- |
| **Vaporized**<br>**Hydrogen Peroxide** | 1 | 8.20 | 1.48 | 6.73 |
|  | 2 | 8.20 | 1.78 | 6.43 |
|  | 3 | 8.20 | 1.48 | 6.73 |

### Validation of N95 respirator filtration efficiency before and after treatment

The filtration validation study conducted as described in the methods section was performed on three new, untreated N95 respirators and on three N95 respirators treated 20 times using the described VHP treatment process. The results of filtration efficiency (see Fig. 3) indicated that at all filtration challenge sizes tested, the measured filtration efficiency of the respirators treated with 20 VHP cycles under the procedures recommended herein remained above the minimum filtration efficiency value of 95% required for the respirators to be classified as an N95 type [15]. The worst-case value of measured filtration efficiency for a 20-cycle treated N95 respirator was found to be 97.89%. These results indicate that the minimum filtration efficiency required for N95 respirator classification is likely to be preserved even beyond 20 treatment cycles.

**Fig. 3.**
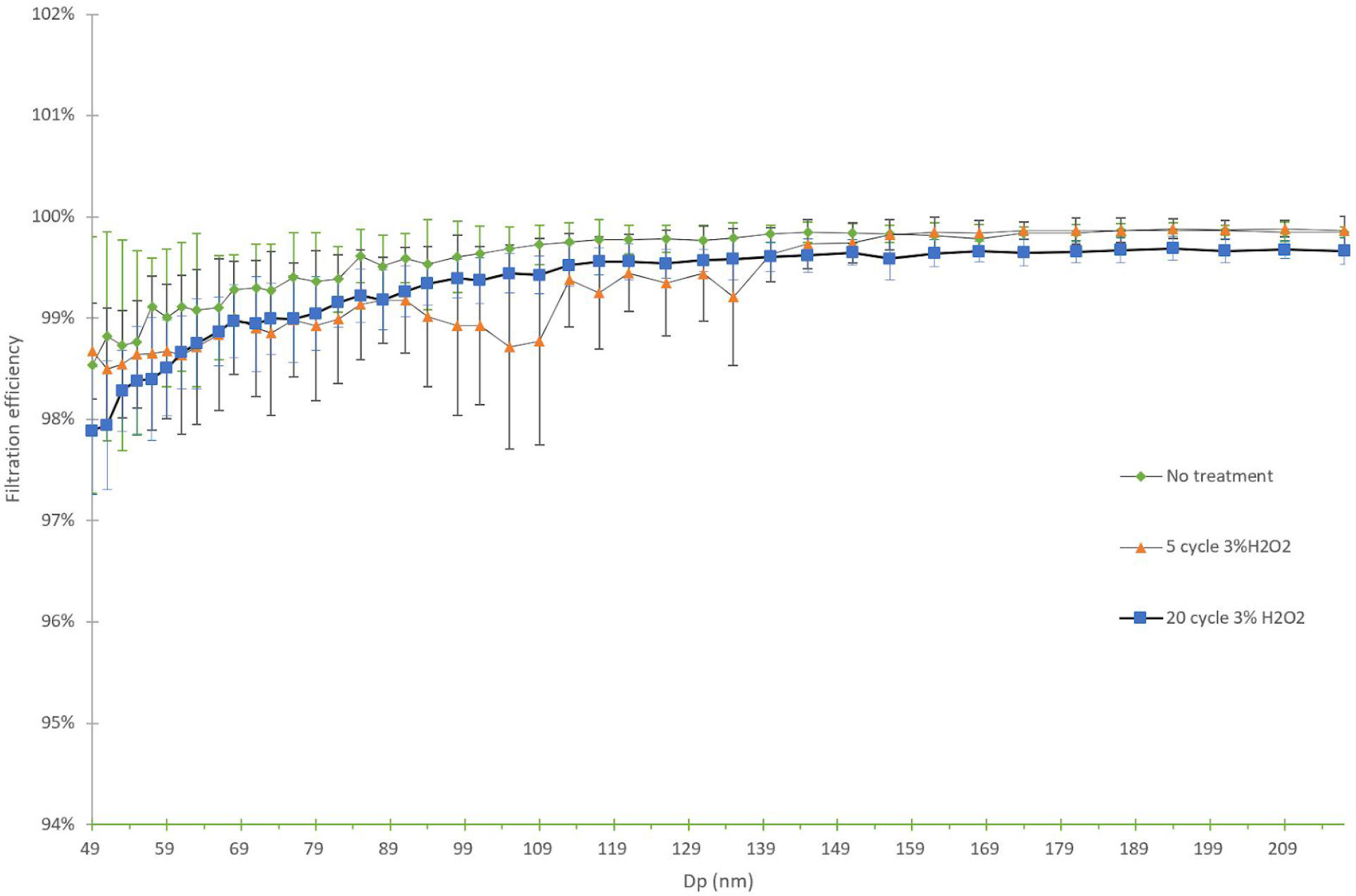
Filtration efficiency of untreated N95 respirators, N95 respirators treated with 5 cycles, and N95 respirators treated with 20 cycles with particle challenge sizes from 50 nm to 300 nm.

### Validation of N95 respirator fluid resistance before and after treatment

The fluid resistance study conducted as described in the methods section yielded the results listed in Table 3. Based on these data, it was shown that the roll-off time increased by an average percent difference of 23% after 5 cycles of VHP treatment. While the ASTM F 1862 standard for testing fluid resistance utilizes synthetic blood at a higher velocity than the droplets used in this non-FDA approved procedure, medical face masks are validated through the FDA by observing that no liquid has penetrated the mask through to the other side [14]. It is notable that no liquid was able to penetrate or absorb through the hydrophobic layer of any of the masks tested. A sample frame of the video capture in which the water drop is about to encounter the mask surface is shown in Fig. 4.

**Table 3.** Results of a water drop deflection test used to quantify performance of the fluid resistant layer on the N95 respirators before and after 5 cycles of VHP treatment.

| **Trial (Experimental):** | **Roll-Off Time (± 17ms):** | **Trial (Control):** | **Roll-Off Time (± 17ms):** |
| --- | --- | --- | --- |
| Mask 1: Drop 1 | 117 | Mask 4: Drop 1 | 83 |
| Mask 1: Drop 2 | 133 | Mask 4: Drop 2 | 100 |
| Mask 1: Drop 3 | 133 | Mask 4: Drop 3 | 100 |
| Mask 2: Drop 1 | 100 | Mask 5: Drop 1 | 100 |
| Mask 2: Drop 2 | 100 | Mask 5: Drop 2 | 83 |
| Mask 2: Drop 3 | 117 | Mask 5: Drop 3 | 100 |
| Mask 3: Drop 1 | 100 | Mask 6: Drop 1 | 83 |
| Mask 3: Drop 2 | 133 | Mask 6: Drop 2 | 100 |
| Mask 3: Drop 3 | 117 | Mask 6: Drop 3 | 83 |
| **Average (ms):** | 117 | **Average (ms):** | 93 |
| **Standard Deviation (ms):** | 14 | **Standard Deviation (ms):** | 9 |
| **Percent Difference:** | 23 |  |  |

**Fig. 4.**
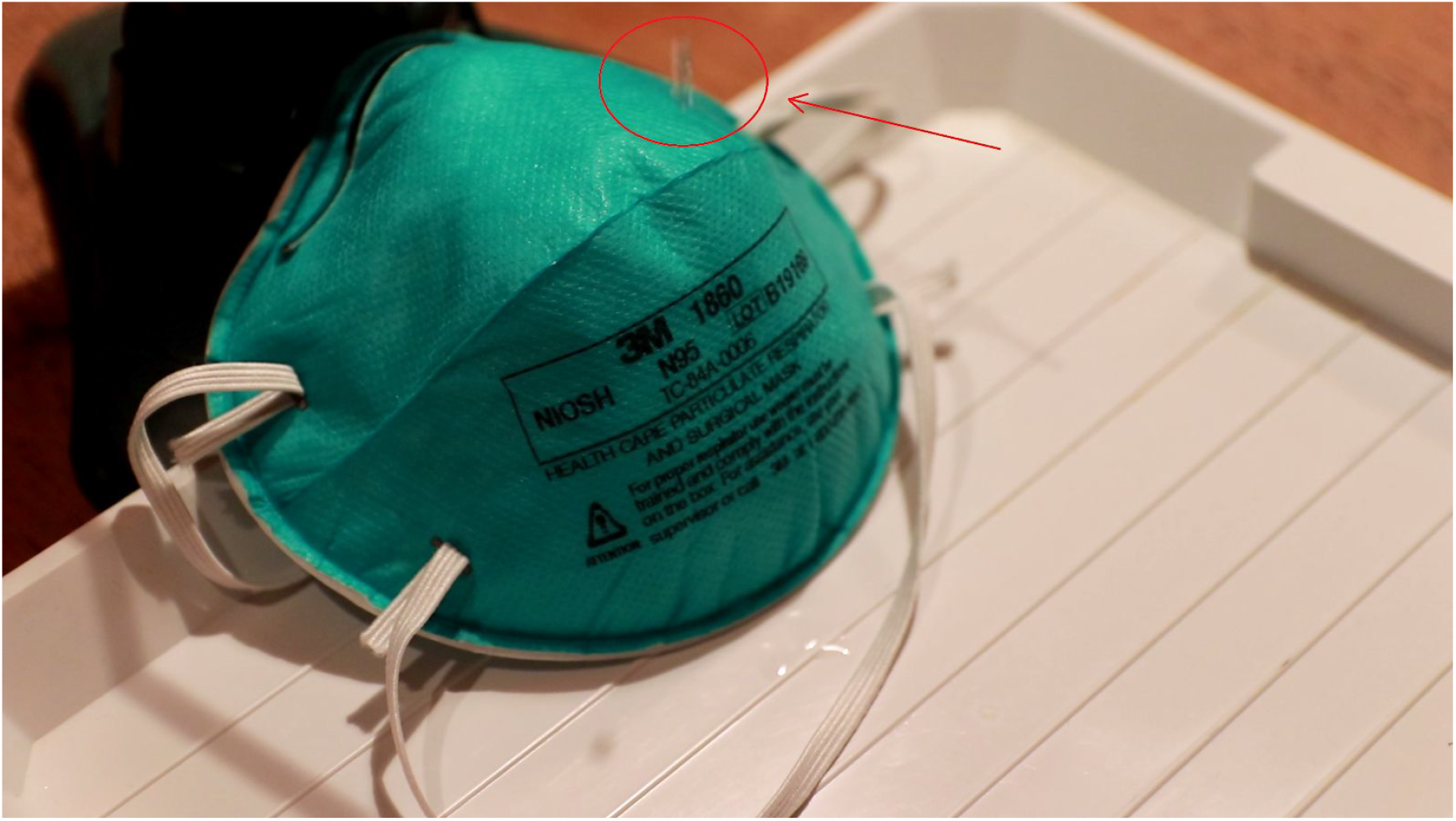
A sample frame of the video capture of the fluid resistance validation test, showing the water droplet prior to encountering the hydrophobic layer of the mask surface.

## Discussion

Common gas-phase disinfectants, such as ethylene oxide (EtO), nitrogen dioxide, peracetic acid, ortho-phthalaldehyde (OPA), glutaraldehyde (Cidex), and ozone, are used to sterilize a variety of healthcare tools and equipment in clinical settings [23]. With the exception of ozone, all of these gases must be obtained as a feedstock material in order to perform sterilization, and many of them possess high toxicity. In particular, ethylene oxide is expensive and known to be carcinogenic and Cidex is an irritant [23]. Furthermore, the required treatment time for EtO is over 12 hours, reducing the throughput of materials through the sterilization system [23]. In contrast, the VHP-based treatment system described in this paper requires only 3% hydrogen peroxide solution, which is inexpensive, widely available, and easy to prepare from higher-concentration solutions if necessary. The byproducts of VHP break down into innocuous water and oxygen [23], and the treatment cycle requires only 60 minutes in the system described in this paper, with an additional 30-minute aeration phase. Combined, these characteristics make VHP a good disinfectant choice for use in both times of crisis and times of normalcy.

The VHP sterilization systems in common use today are large, high-volume systems that are most suitable to large healthcare facilities where both capital and supporting infrastructure are available. For example, one commonly-employed VHP system is the Bioquell BQ-50 hydrogen peroxide vaporization system. This system is costly, designed to operate in a large dedicated room on the order of 200 cubic meters, and requires a feedstock of 35% hydrogen peroxide solution to operate [10]. Such systems are not well-suited to smaller healthcare sites such as urgent care clinics or assisted living facilities, nor are they accessible to commercial partners or small businesses. The VHP system described in this paper occupies a space less than 1 cubic meter and only requires a standard 3% hydrogen peroxide solution as feedstock. It can also be scaled up to larger installations as needed, simply by using additional units operated in parallel. As such, we believe that the VHP system described in this paper will provide smaller healthcare facilities and other businesses with unprecedented access to easy-to-use, affordable sterilization technology.

To verify the performance of this device, the system’s virucidal efficacy, impact on the filtration efficiency of N95 respirators, and compatibility with fluid-resistant N95 respirators were assessed experimentally. In regard to virucidal efficacy, the concentration of VHP in the test chamber was characterized with commercially available hydrogen peroxide test strips under the recommended VHP chamber operating conditions. This test indicated that an operational VHP concentration of 573 ± 107 ppm was achieved in the chamber. This VHP range, 466 ppm - 681 ppm, significantly differed from the analytically-derived target concentration of 1200 ppm within the chamber. This disparity can be attributed to several possible causes. In particular, insufficient vacuum level, changes in the surface temperature of the nonreactive fiber matrix (NFM) during vaporization, and potential leaks within the vacuum chamber could have resulted in this disparity.

Despite this difference, the observed reduction in viral load indicates that the process described herein is effective in sterilizing N95 respirators even at the reduced VHP concentrations measured experimentally. In fact, an exposure time of 60 minutes in recommended chamber operating conditions was shown to provide greater than 6-log_10_ reduction in the P22 bacteriophage used as a surrogate for SARS-CoV-2. This result not only meets but exceeds the ATSM definition for sterilization [22].

While the sterilization of N95 respirators with a scalable VHP system represents a consequential achievement, the impact would be mitigated if the integrity of the respirator was compromised. However, N95 respirators treated with 20 cycles of 60-minute exposure in the recommended chamber operating conditions remained above the minimum required filtration efficiency of 95% for this type of respirator, indicating that the VHP treatment process does not pose a significant risk to filtration efficiency in the concentrations, time, and number of cycles studied. Similarly, the observed 23% average percent increase in water drop roll-off time after 5 cycles of VHP treatment, combined with the lack of visible liquid penetration, indicates that the recommended VHP conditions did not render the hydrophobic layer unusable. While fit and inhalation resistance testing of the masks before and after treatment were not performed in this study, existing literature suggests that VHP has no detrimental effect on either [4, 5].

It is important to note that vaporized hydrogen peroxide must be properly destroyed or dispersed before PPE is donned by personnel. NIOSH has established a recommended exposure limit of no greater than 1 ppm time-weighted average for vaporized hydrogen peroxide exposure [24]. Specific guidelines for minimizing exposure of personnel to high hydrogen peroxide levels are addressed in the instructions for use provided with the hydrogen peroxide treatment system (see Supporting Information). In particular, it is recommended that the operator keep their face away from the VHP treatment chamber during opening and removal of items, and that the VHP treatment chamber be opened in a well-ventilated space. Prior to removing items, the watch glass of hydrogen peroxide solution should be removed from the chamber, the mask loader and items being sterilized should be placed back into the chamber, and the vacuum pump should be run continuously for 30 minutes to evacuate the chamber and remove the remaining VHP from the mask surfaces prior to the removal of masks from the chamber. The system should not be operated if there appears to be any flaw in its construction or visible damage.

As an additional safety precaution, the suggested strength of hydrogen peroxide solution to be used with this system is 3% due to its relatively low concentration. While any solution of hydrogen peroxide can act as an oxidizing agent and should be handled with care as specified by H_2_O_2_’s material safety data sheet [25], a concentration of 8% or greater of hydrogen peroxide is considered an oxidizer [26]. Using a solution with a concentration of 8% or greater requires more intensive safety measures than outlined in the guidelines for operation of our VHP sterilization system. As an added benefit, 3% hydrogen peroxide solution is also widely available.

Finally, while this work found no significant effect on N95 respirator filtration efficiency after 20 cycles of treatment and only minor effect on fluid resistance of N95s after 5 cycles treatment, consideration must be given to the real world application of a VHP system for N95s. With extended use of N95 respirators during epidemic-related shortages, respirators can experience significant wear and outside debris contamination that limit N95 performance and fit. A decontaminated mask should be inspected before wearing. Sterilized masks should be returned to the original user to prevent adverse effects on fit and performance. Precautions against wear of use on N95 respirators, such as reusing them for less than the validated 20 cycles, should be taken, especially in healthcare settings with extended use of respirators. Further precautions are outlined in the “Considerations for the Safe Reuse of N95 Respirators” section of Supporting Information.

## Conclusion

Achieving a 6-log_10_ reduction in P22 bacteriophage viral loads suggests that our VHP system will be effective in eliminating SARS-CoV-2 on N95 respirators. Furthermore, decontaminated N95 respirators showed no significant decrease in standard filtration performance when compared to untreated N95 respirators. Along with existing literature supporting VHP’s ability to safely decontaminate N95s, these data demonstrate the capability of this system to effectively sterilize standard N95 respirators and face coverings without a reduction in performance.

While developed and validated for clinical application, the low cost and ease in manufacturing of this system makes it scalable to the needs of businesses operating during the COVID-19 epidemic. Our VHP system can be used by barber shops, schools, and restaurant employees, among a myriad of other applications, to safely sterilize N95 respirators, polypropylene-based masks, and other face coverings for reuse. In addition to respirators, this system may also be suitable for use on hard materials typically found in household and commercial settings, such as tools, office supplies, and other frequently-handled items.

Allowing businesses to safely reuse face coverings and other PPE will have numerous economic and societal benefits. First, this technology will encourage businesses to follow public health guidelines by reducing the financial burden of compliance. While the cost of masks and PPE may seem small, they quickly become significant if each employee is provided with a new face covering daily. However, reusing these coverings will make PPE purchases more infrequent and feasible. This will quickly offset the costs of the sterilization device and can even allow businesses to invest in higher quality PPE when it becomes available.

Helping businesses reuse PPE will also have the added benefit of decreasing overall demand for these devices. This will mitigate the strain on existing PPE supply chains even as increasingly large sectors of the economy resume in-person operations. As a result, this will help ensure that basic PPE remains available throughout the future progression of the COVID-19 pandemic.

Furthermore, incorrect reuse of contaminated facial coverings can decrease efficacy and even act as a vector for viral transmission. Implementation of these VHP treatment systems will give business owners access to technologies never before implemented outside of medical facilities, allowing them to process PPE in the same way it would be treated in a healthcare setting. Thus, business owners can implement re-use of facial coverings with confidence, knowing they are protecting the health of their community as they resume operations.

Finally, the implementation of sterilization devices will inspire consumer confidence as communities resume in-person economic activity. This will be facilitated by the widespread availability of safe, quality facial coverings and PPE to business owners, their employees, and their customers. In turn, consumers will feel safer in these places of business, and will therefore more regularly support businesses,bolstering the newly reopened economy. Taken together, this work highlights the efficacy of our novel, scalable, and cost-effective VHP system, which can be rapidly deployed for both clinical and community applications.

## Data Availability

All raw data is available upon request.

## Acknowledgements

Capture efficiency testing was supported by the NSF RAPID program (CBET-2028074). Other funding support was provided by the Arizona State University Knowledge Enterprise and the NSF Water and Environmental Technology (WET) Center at Arizona State University (award number 1361815).

## Supporting Information

### System Construction

The mask carrier structure is assembled from a subset of the components listed in Table

Four 0.25 inch holes are drilled in one of the perforated metal dishes, positioned so that three holes are arranged equidistant at 120 degrees along the outer lip of the first perforated dish, and one hole is located at the center of the disc. One centrally-located 0.25 inch hole is drilled in each of the two remaining perforated metal discs. A 2 inch bolt is inserted through each of the three equilaterally-spaced holes in the first disc. A washer is placed on each side of the perforated disc material to evenly distribute the mounting force, and nylock nuts are used to secure the 2 inch bolts to the first disc. These three bolts are the feet of the structure and provide support to the rest of the structure. An acorn nut is tightened onto the end of the 8 inch all-thread bolt, which serves to confine rotation of the bolt. The all-thread bolt is inserted through the central holes in the three perforated discs, washers are placed on either side of each disc, and nylock nuts are used to secure each disc in place on the 8 inch all-thread bolt. The perforated discs are vertically arranged approximately 6.5 cm apart. A final nylock nut is threaded onto the top of the all-thread bolt to serve as a handle for removing the mask loader structure. The finished mask loader structure appears as depicted in Fig. 5.

**Fig. 5.**
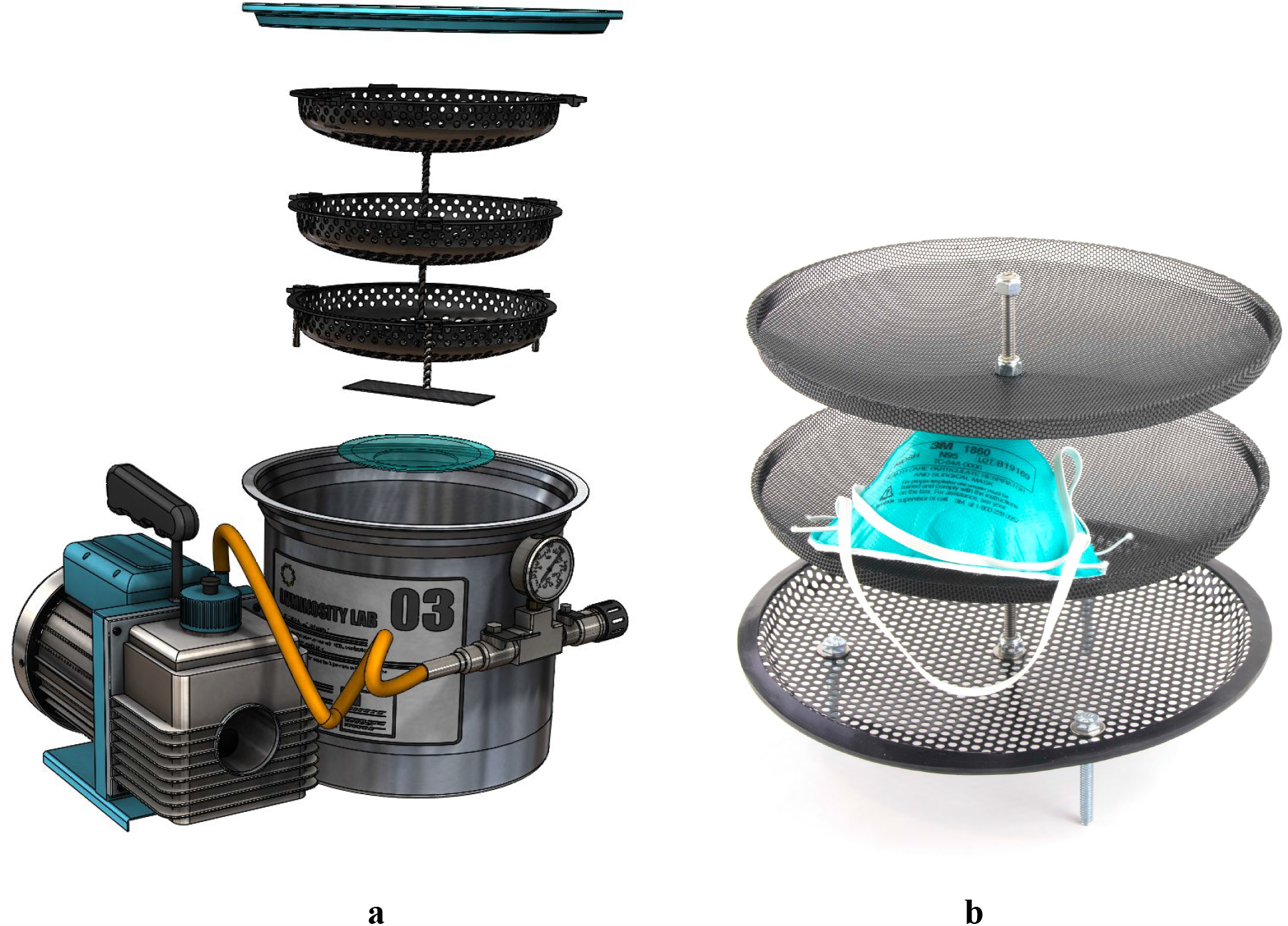
a. A digital rendering of the vacuum pump and vacuum chamber with glass lid, hydrogen peroxide dish, and mask loader tray. b. A photograph of a mask loader tray that can fit up to 12 N95 respirators, consisting of three perforated metal dividing plates to increase diffusion of VHP into the respirators.

The vacuum system is assembled from a subset of the components listed in Table 4. After verifying the oil level in the vacuum pump, the vacuum pump is connected to the vacuum chamber using the flare-fitting hose provided with the vacuum chamber. The provided air intake filter is threaded onto the opposite side of the vacuum chamber intake manifold. Finally, the rubber of the vacuum gauge oil plug is removed to ensure that the vacuum gauge can equilibrate with the local atmospheric pressure.

**Table 4.** Bill of materials and associated costs of our VHP sterilization system.

| Component: | Quantity | Approximate Cost (Per Component): |
| --- | --- | --- |
| Vacuum Chamber (5 Gal) | 1 | \$245.00 |
| Rotary Vane Vacuum Pump | 1 | \$54.99 |
| Perforated Metal Disc | 3 | \$4.45 |
| 8" All-Thread Bolt ($\frac{1}{4}$" ) | 1 | \$0.86 |
| 2" Bolt ($\frac{1}{4}$" ) | 3 | \$0.14 |
| Washer ($\frac{1}{4}$" ) | 12 | \$0.04 |
| Acorn Nut ($\frac{1}{4}$" ) | 1 | \$0.09 |
| Nylock Nut ($\frac{1}{4}$" ) | 9 | \$0.05 |
| Watch Glass | 1 | \$1.50 |
| Fiberglass Patch (Consumable) | 5 cm x 15 cm<br>(1 cycle) | \$0.01 |
| 3% H<sub>2</sub>O<sub>2</sub> Solution (Consumable) | 10 ml/cycle | \$0.01 |
| Vacuum Pump Oil (Consumable) | 16 ml/cycle | \$0.16 |
| | **Total Cost:** | **\$317.32** |

The nonreactive fiber matrix (NFM) is assembled from a subset of the components listed in Table 4. A 5 cm by 15 cm strip of fiberglass cloth is cut from the provided roll of fiberglass. This strip, which serves as the NFM, is tri-folded so that the folded patch measures 5-cm by 5-cm and is 3 layers thick. The NFM is then placed into a watch glass, which is positioned at the bottom center of the vacuum chamber.

When the system is ready to operate, 10 ml of 3% hydrogen peroxide solution is dispensed into the watch glass. A stir-rod or wood splint is used to tamp down the NFM and to saturate it in hydrogen peroxide solution. An example of a prepared watch glass and NFM is shown in Fig. 6 (outside the vacuum chamber).

**Fig. 6.**
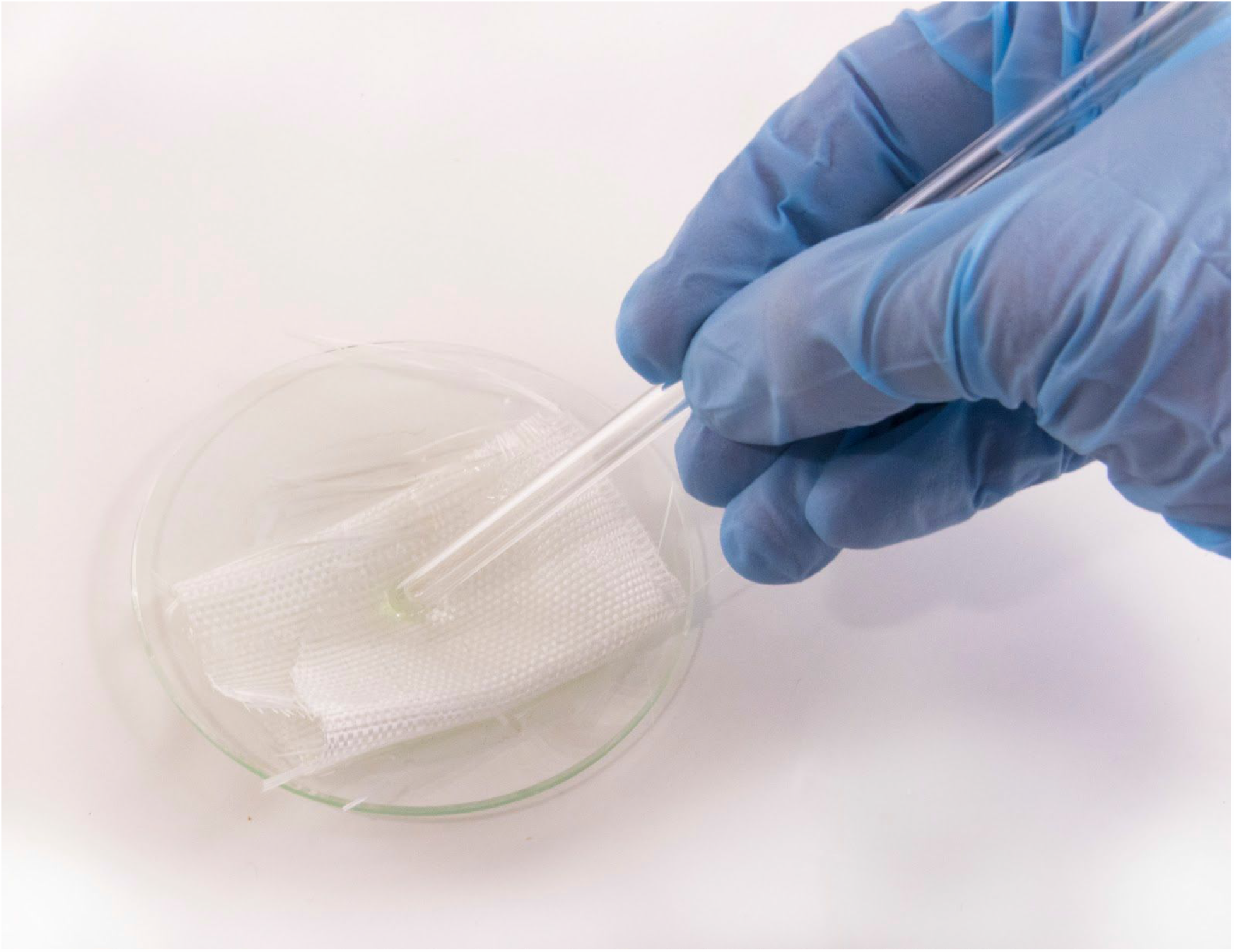
A woven fiberglass cloth swatch, used as a nonreactive fiber matrix to increase the rate of vaporization, placed into a dish of hydrogen peroxide solution. The action of soaking the cloth in hydrogen peroxide solution by use of a stir rod is depicted.

### Dosing for Sterilization

The dosage required for the VHP system to reach is dependent on the device being decontaminated and what sterility assurance level is required. N95 respirators should be sterilized due to their proximity to the respiratory tract of health care professionals. According to the American National Standard ANSI/AAMI ST67, a 6-log_10_ reduction of microorganisms is required for sterilization of the majority of medical devices [22], which is extended to N95 respirators. Table 5 shows the dosage recommendations for achieving sterilization using our VHP system.

**Table 5.** Dosage chart containing pressure level and phase times in order to reach a 6-log10 reduction of biological contaminants.

|  | **Suggested Use Cases** | **Hg Achieved** | **Gassing Phase\*** | **Dwell Phase\*\*** | **Aeration Phase\*\*\*** |
| --- | --- | --- | --- | --- | --- |
| **Sterilization (6-log<sub>10</sub> reduction)** | N95 Masks and other filtering facepiece respirators, surgical masks, other PPE (specifically required for healthcare facilities) | Vacuum level indicated by (1) | 3-5 min (until vacuum level indicated by (1) reached) | 60 min | 15 min |

\*The gassing phase is the time required for the chamber to reach the recommended pressure.
\*\*The dwell phase indicates the length of VHP contact suggested for N95 masks and other use cases when the chamber is left undisturbed.
\*\*\*The aeration phase refers to the process of reintroducing atmospheric air into the chamber to prevent off-gassing from PPE during use.

### System Operation

The vaporized hydrogen peroxide system is operated as follows. First, masks or items to be treated are placed onto the three layers of the mask loader structure within the vacuum chamber with the seal facing downwards, flat on the tray, and the largest area of the respirator faced upwards. If possible, masks should be placed into self-sealing sterilization pouches before placement in the VHP system to prevent decontamination after treatment. The NFM and hydrogen peroxide solution are prepared as described above. The populated mask loader structure is lowered into the vacuum chamber, above the watch glass with the NFM. The silicone-gasketed glass lid provided with the vacuum chamber is placed onto the top of the vacuum chamber, and it is ensured that the silicone gasket that surrounds the glass lid is uniform all around the lid and is not buckled or loose in any region. It is also verified that the glass lid is centered on the chamber. At this point the atmospheric admission valve (the rightmost valve with the air filter connected to the vacuum chamber) is closed and the vacuum valve (the leftmost valve connected to the vacuum pump via the hose) is opened. The vacuum pump is turned on, and 3-5 minutes are waited until the vacuum gauge has reached the vacuum level indicated by equation (1). The terminal vacuum level will depend on the ambient pressure (altitude), and may need to be corrected for higher or lower altitude facilities. The vacuum pump isolation valve is closed once the terminal pressure is achieved, and the vacuum pump is turned off. Once treatment has begun, the chamber is allowed to stand undisturbed for 60 minutes in order to achieve a dose of VHP exposure recommended in Table 5. Orienting the N95s within the chamber so the largest area of the respirator faces upward allows for maximum exposure to VHP and higher quality disinfection [28]. After the treatment period is complete, atmospheric air is re-introduced using the atmospheric admission valve and the lid is removed from the vacuum chamber. The mask loader structure is taken out of the chamber and the hydrogen peroxide watch glass is removed. The mask loader is then re-inserted into the chamber, the atmosphere valve is closed, and the vacuum pump is switched back on. The vacuum pump is allowed to run continuously for 30 minutes to ensure the bulk removal of hydrogen peroxide and other volatile materials. Finally, the atmosphere valve is again opened to re-pressurize the chamber by removing the lid, and treated items may be removed from the mask loader structure.

After treatment is complete, relevant disposal guidelines are followed to ensure safe and environmentally conscientious treatment of waste products generated by the system. The watch glass with the NFM and hydrogen peroxide solution in it is removed from the chamber. The NFM is removed from the watch glass and disposed of in a hydrogen peroxide-compatible chemical solid waste collection process. Any remaining hydrogen peroxide solution is disposed of in a peroxide-compatible chemical liquid waste collection process. The watch glass is rinsed with water and left to air-dry. The watch glass may be reused in subsequent treatment cycles.

The system should be operated at approximately 28 torr absolute pressure. The relative pressure indicated by the vacuum gauge (in inches of Hg) is computed as a function of the ambient pressure in the geographic region of operation (in mmHg) in (8):

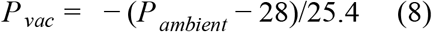

where *P*_*vac*_ is the relative pressure that must be achieved in the vacuum chamber in inches of Hg and *P*_*ambient*_ is the barometric pressure of the region in which the system is being operated, in mm of Hg. *P*_*ambient*_ can be obtained from a standard weather report in the region where the system is operated. After every 30 treatment cycles, vacuum pump oil should be replaced with automotive vacuum pump oil to maintain pump performance.

### Aeration Phase

The following discussion provides justification for an aeration phase of fifteen minutes to prevent personnel exposure to VHP. According to *N95 Mask Decontamination using Standard Hospital Sterilization Technologies*, the necessary aeration time for N95 masks treated with VHP at atmospheric pressure at 750 ppm concentration is 20 minutes [8]. The proposed VHP system employs a vacuum drying stage that operates at 28 torr. The proposed system also utilizes a concentration that is estimated to reach a maximum value of 681 ppm by experimental measurement.

If the rate of evaporation of hydrogen peroxide from surfaces is approximated to be linearly dependent on ambient pressure, the required aeration time can be expressed in terms of the ratio of ambient pressures and the original aeration time in (9):

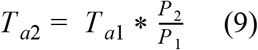

where *T*_*a*2_ is the aeration time at ambient pressure *P* _2_ and *Ta*1 is the aeration time at ambient pressure *P* _1_.

Furthermore, if the aeration time is assumed to be approximately linearly dependent on the concentration of VHP during treatment, the required aeration time can be expressed in terms of the ratio of treatment concentrations and the original aeration time in (10):

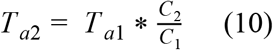

where *T*_*a*2_ *C*_1_. is the aeration time at concentration *C*_2_ and *Ta*1 is the aeration time at concentration

Combining these equations accordingly, the aeration time as a function of both the ambient pressure ratios and concentration ratios between the proposed VHP system and existing VHP systems in the literature can be expressed as in (11):

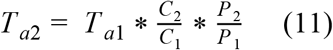

Substituting the given values of concentration and pressure, the required aeration time in the vacuum drying process is therefore expressed in (12)

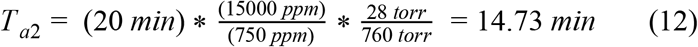

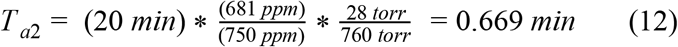

It requires approximately 5 minutes of vacuum time to achieve the vacuuming time to achieve the vacuum level under which this aeration time was computed using the pumps recommended in the bill of materials for this system. The total minimum vacuum pump operation time during the aeration phase is therefore approximately 5.67 minutes. Due to tolerances in the vacuum pump design and chamber temperature, a conservative recommendation for aeration time can be given as twice this value plus a margin of error, for a total recommended aeration vacuum pumping time of 15 minutes.

### Considerations for the Safe Reuse of N95 Respirators

Certain precautions should be taken when reusing N95 respirators. The CDC recommends reuse of disposable N95s only during times of shortage [9]. With extended use of N95 respirators during epidemic-related shortages, respirators can experience significant wear and outside debris contamination that limit N95 performance and fit and reduce the ability of an N95 to be reused safely. With any sterilization and reuse of N95s, significant attention should be given to donning and doffing procedures to prevent contamination of healthcare personnel. A system for returning the respirators back to the original health care workers should be put in place, as outlined in reference 28. It is highly suggested to track the N95s by marking their containers to ensure each respirator has a single user, which reduces concerns about fit of the mask or complications such as the original user wearing cosmetics or lotion [28]. While a user seal check is required in many healthcare and industrial facilities, it is highly recommended that all personnel using a sterilized respirator should conduct one. At any visible deformation of shape, debris, or contaminate on the N95, the respirator should be disposed of [28]. Measures such as placing the respirators in self-sealing sterilization pouches, if possible, prevent viral re-contamination during transportation after treatment [28].

## Notes

### Competing Interest Statement

The authors have declared no competing interest.

### Author Declarations

No human or animal subjects were used in this work.

